# Variability in First Academic Medicine Job Offers in Pulmonary and Critical Care Medicine

**DOI:** 10.1101/2025.09.05.25334955

**Authors:** Emily M Olson, Matthew E Modes, Timothy J Rowe, Patrick G Lyons, Nicholas E Ingraham, Nandita R Nadig, Clara J Schroedl, Catherine A Gao

## Abstract

**Background:** A minority of Pulmonary and Critical Care Medicine (PCCM) graduates pursue careers in academic medicine. Although compensation is only a portion of the career decision, job negotiations remain shrouded in ambiguity and inconsistency. Additionally, while role-level salary tables exist through the Association of American Medical Colleges (AAMC), to our knowledge there is no resource that includes important non-salary information such as start-up packages, full-time equivalent (FTE) breakdown, and bonus ranges.

**Objective:** We sought to describe key components of first PCCM academic job offers for both physician scientists and clinician educators, including salary, start-up packages, non-clinical FTE, and bonuses.

**Methods:** An electronic survey was distributed via a snowball method between May - June 2025. PCCM graduates between 2020-2025 who accepted a job in academic medicine were included. Mann-Whitney Wilcoxon tests were used for ordinal comparisons. Qualitative analysis of free text responses was performed with a social cognitive career theory framework.

**Results:** There were 60 respondents who provided information about 103 job offers, with 50% (14/28) of physician-scientists and 66% (21/32) of clinician-educators reporting more than one job offer. Physician-scientists received lower salary offers compared to clinician-educators (respective median ranges: $150,000-$199,999 vs $250,000-$299,999, p<0.001). 35.7% physician-scientists (10/28) received a career development award prior to negotiation, which was associated with a higher start-up package offer (p<0.05). For all clinician-educator jobs (n=59), 42.4% had non-clinical FTE in the initial offer. Many respondents commented on the lack of negotiating power.

**Conclusion:** PCCM physician-scientists and clinician-educators experience wide variability in their initial job offers. Recognizing differences is essential to improve transparency in job negotiations in academic medicine.

## Introduction

The transition from training to a faculty position in academic medicine is a critical juncture, and often driven by external factors (such as location, work/life balance, family) as much as career goals.(1, 2) In Pulmonary and Critical Care Medicine (PCCM), a minority of graduates choose careers in academic medicine.(3) Additionally, even fewer pursue research-focused careers.(4) Despite its importance, the process of negotiating job offers - including salary, startup packages, and clinical full-time equivalent (FTE) - remains shrouded in ambiguity and inconsistency. Negotiations are often conducted with limited transparency regarding the terms and expectations of academic positions.(5, 6) Additionally, there has been significant concern about lower salaries and academic support for faculty members identifying as women or underrepresented in medicine.(7–11) The lack of standardized practices not only creates disparities in compensation and non-monetary support, but also hinders the broader dialogue about what constitutes fair and competitive academic employment.

While region and role-level salary tables exist through the AAMC (Association of American Medical Colleges) and the AMA (American Medical Association) databases, many require payment or memberships, and may not be accessible to all graduates.(12) Other online databases like Medscape or Doximity report an average salary but do not stratify by practice settings.(13, 14) This is a key distinction as academic physicians often earn less compared to private practice.(15) Additionally, these resources do not include important non-salary information such as clinical FTE, promotion pathway details, or startup packages. All of these provide important context to help graduates understand the market for their skills and expertise.

To our knowledge, there are no prior studies describing key components of first PCCM job negotiations in academic medicine. We sought to address this gap by exploring the range of job offers received by early career PCCM physician-scientists and clinician-educators in an exploratory study. We aimed to compile a reference that can help improve equity during job searches.

## Methods

A survey was developed to capture information about demographics, key components of faculty offers (salary, clinical FTE, startup packages), and the results of negotiation (**Supplemental Material 1. Survey**). The survey instrument was designed based on a review of relevant literature about challenges in recruiting to academic medicine, including finances and protected time.(5, 16–22) CAG, NEI, PGL, JAH, PDLC, MEM, and NRN used their expertise as physician-scientists and EMO, TJR, SAL, and CJS provided expertise as clinician-educators to write the survey. After pilot testing with eight early career faculty, iterative revisions were made to ensure clarity, face validity, and comprehensiveness.(23) Respondents had the opportunity to answer questions for up to three job offers, meaning they could answer between 28 and 85 individual questions depending on their identified career. The Northwestern University IRB reviewed the study questions, design, and consent language and deemed this study exempt (STU00223858).

The final survey was administered electronically using a secure, web-based survey platform, REDCap,(24) between May and June 2025. Invitations to participate were distributed via personal email, in-person QR codes, and social media platforms (Twitter/X and Bluesky). PCCM faculty who graduated between 2020-2025 and accepted a job in academic medicine were eligible for inclusion. Participation was voluntary, and informed consent was obtained from all respondents prior to survey commencement. We chose to distribute the survey via a snowball sampling technique due to the private nature of discussing finances and the relatively small number of recent PCCM graduates entering academic medicine. Snowball sampling is frequently used in exploratory research when wanting to recruit individuals with specific characteristics.(25) This survey distribution method is particularly helpful when there is no existing database and when the pre-existing relationship with contacts may encourage participants to share experiences. Prior surveys assessing perceived gender disparities on career advancement had similar success with this technique.(26)

Descriptive statistics were calculated and presented using median (Quartile 1, Quartile 3) and n(%). Comparisons were performed based on prespecified hypotheses that there would be differences based on K status, gender and staying at institution of training. Mann-Whitney Wilcoxon tests were used for comparisons between ordinal data such as salary, with Bonferroni correction for multiple comparisons. All analyses were performed using Python 3.9.6 with the following packages: pandas 2.2.3, matplotlib 3.9.3, seaborn 0.13.2, scipy 1.13.1, tableone 0.9.5, statannotations 0.7.2, and numpy 2.0.2. The code for our analysis is available at: https://github.com/cag-lab/pccm_job_negotiation.

Qualitative analysis of open-ended text box responses was performed by researchers with qualitative training (MEM and EMO) using a general inductive approach with a framework of social cognitive career theory.(27) Comments were read multiple times through an iterative process to summarize the findings. No software was used.

## Results

We received 60 responses. We had directly contacted 94 individuals about the survey. Our response rate is at the highest, 64% (60/94) and likely lower given the snowball sampling method and an unknown true denominator. Of the 60 respondents, 28 identified as physician-scientists (47%) and 32 as clinician-educators (53%) (**Table 1**). Physician-scientists represented a range of research areas: 21.4% basic science, 21.4% translational, 75% clinical, and 21.4% computational (respondents were allowed to select more than one research area).

**Table 1:** Respondent demographics, broken down by physician-scientist and clinician-educators (n(%)). URIM = underrepresented in medicine.

|  | All Respondents<br>(n=60) | Physician-Scientist<br>(n=28) | Clinician-Educator<br>(n=32) |
| --- | --- | --- | --- |
| *Hired at Home Institution* | 34 (36.7) | 20 (71.4) | 14 (43.8) |
| *Multiple Offers* | 37 (61.7) | 14 (50.0) | 23 (71.9) |
| *Additional Degree(s)* |  |  |  |
| MS, MPH, MSCI, etc. | 28 (46.7) | 20 (71.4) | 8 (25.0) |
| PhD | 2 (3.3) | 2 (7.1) | 0 |
| *Gender* |  |  |  |
| Woman | 34 (56.7) | 12 (42.9) | 22 (68.8) |
| Man | 25 (41.7) | 15 (53.6) | 10 (31.2) |
| Prefer not to answer | 1 (1.7) | 1 (3.6) | 0 (0.0) |
| *Race/Ethnicity* |  |  |  |
| White | 42 (70.0) | 18 (64.3) | 24 (75.0) |
| Asian or Asian American | 13 (21.7) | 7 (25.0) | 6 (18.8) |
| Black or African American | 2 (3.3) | 0 (0.0) | 2 (6.2) |
| Hispanic, Latino, Latina, or LatinX | 3 (5.0) | 3 (10.7) | 0 (0.0) |
| Prefer not to Answer | 1 (1.7) | 1 (3.6) | 0 (0.0) |
| *Self-Identifies as URIM* | 6 (10.0) | 3 (10.7) | 3 (9.4) |

There were a total of 103 job offers, with 33 respondents providing information on multiple offers. Missing responses amounts are reported in the **Supplemental Data 1**. 50.0% (14/28) of physician-scientists and 65.6% (21/32) of clinician-educators reported more than one job offer. Ten physician-scientists and fifteen clinician-educators had two offers; three physician-scientists and six clinician-educators had three offers.

The median salary range for physician-scientists was $150,000-$199,999 (Q1: $150,000-$199,999, Q3:$200,000-$249,999), compared to $250,000-$299,999 (Q1:$200,000-$249,999, Q3:>$300,000) for clinician-educators (p<0.001). There was a significant difference between receiving the offer from the institution where they trained (p<0.01), if the respondent had multiple job offers (p<0.001), and cost of living between high (such as New York, NY) and low (such as Wichita, KS) cities (p<0.01) (**Figure 1**). There were no differences in salary based on gender. For 40% (22/55) clinician-educator job offers, respondents indicated the institution having transparency in salary equity. There was salary adjustment for RVUs in 49.1% (27/55) of clinician-educator job offers. 45.5% of physician-scientists received a bonus and 55.9% of clinician-educators received a bonus. A subset of respondents did not know if a non-compete clause existed for specific job offers (31.8% of physician-scientists and 23.7% clinician-educators). Non-compete clauses are described in the **Supplemental Table 1**.

**Figure 1.**
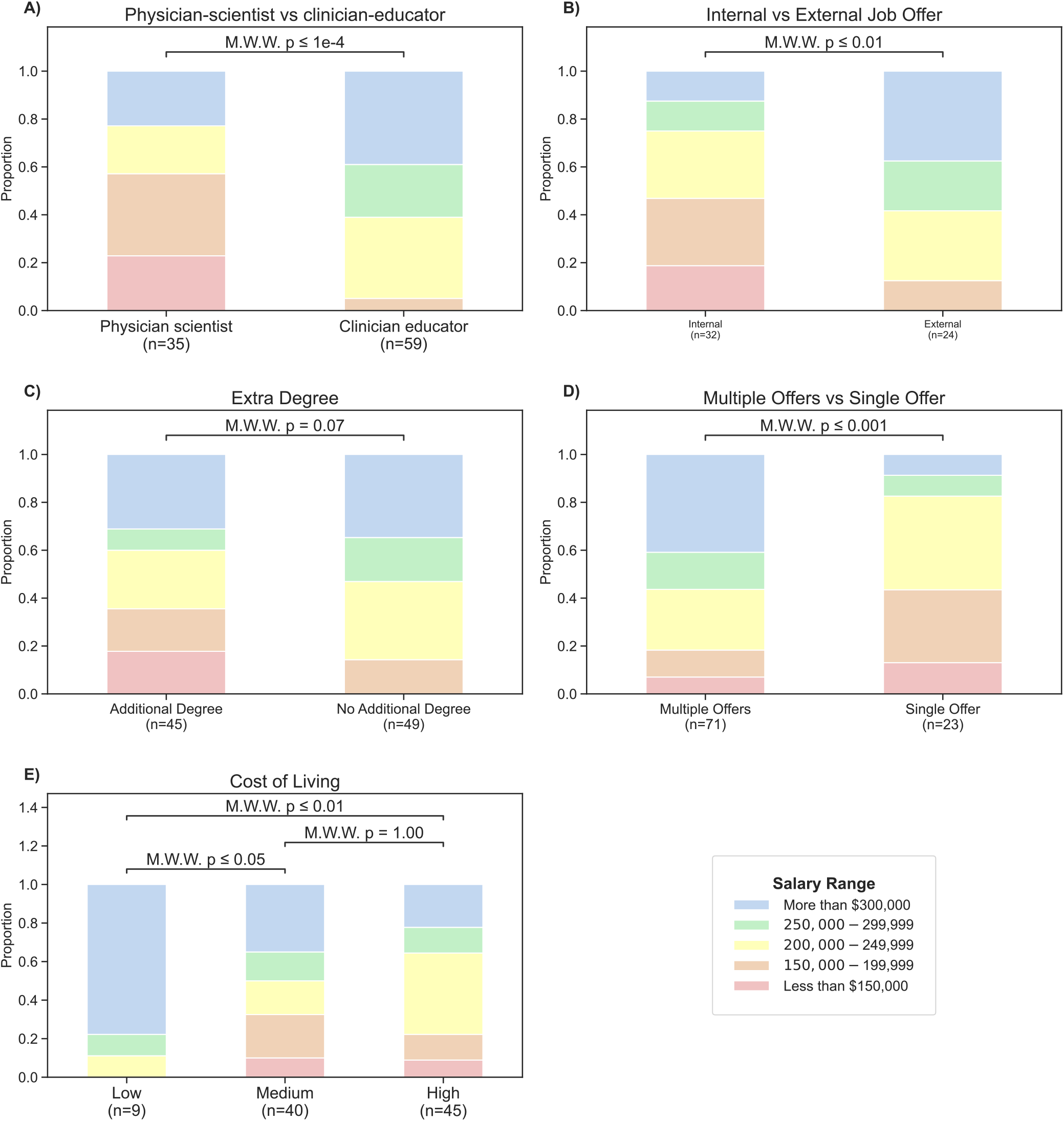
Proportional salary range distribution comparisons for all job offers (n=96 reported salary amounts). Sample size below each bar. Statistical comparisons performed using Mann-Whitney Wilcoxon (M.W.W.) test with p-values shown above bars. (A) Physician-scientists versus clinician-educators. (B) Internal versus external job offers. (C) Physicians with additional advanced degrees (MS, PhD, etc.) versus MD only. (D) Recipients of multiple offers versus single offer. (E) Estimated cost-of living.

Physician-scientists provided details about 44 job offers (**Table 2**). Ten physician-scientists (35.7%, 10/28) had received an early career development award (CDA, such as a K08 or K23) prior to job negotiation. Having a CDA was not associated with salary. Over half of job offers (54.5%, 24/44) included a startup package. Nine offers (20.5%) had cost-sharing, which was described as the grant not covering all the salary and the individual needing to pay the differential via startup funds. Higher startup packages were offered to respondents who had a CDA prior to negotiation (p<0.05), identified as women (p<0.05), or included salary cost-sharing (p<0.05) (**Figure 2**). There was no significant difference in the startup package if the respondent had multiple offers, extra degrees, or between internal/external offers

**Table 2:**
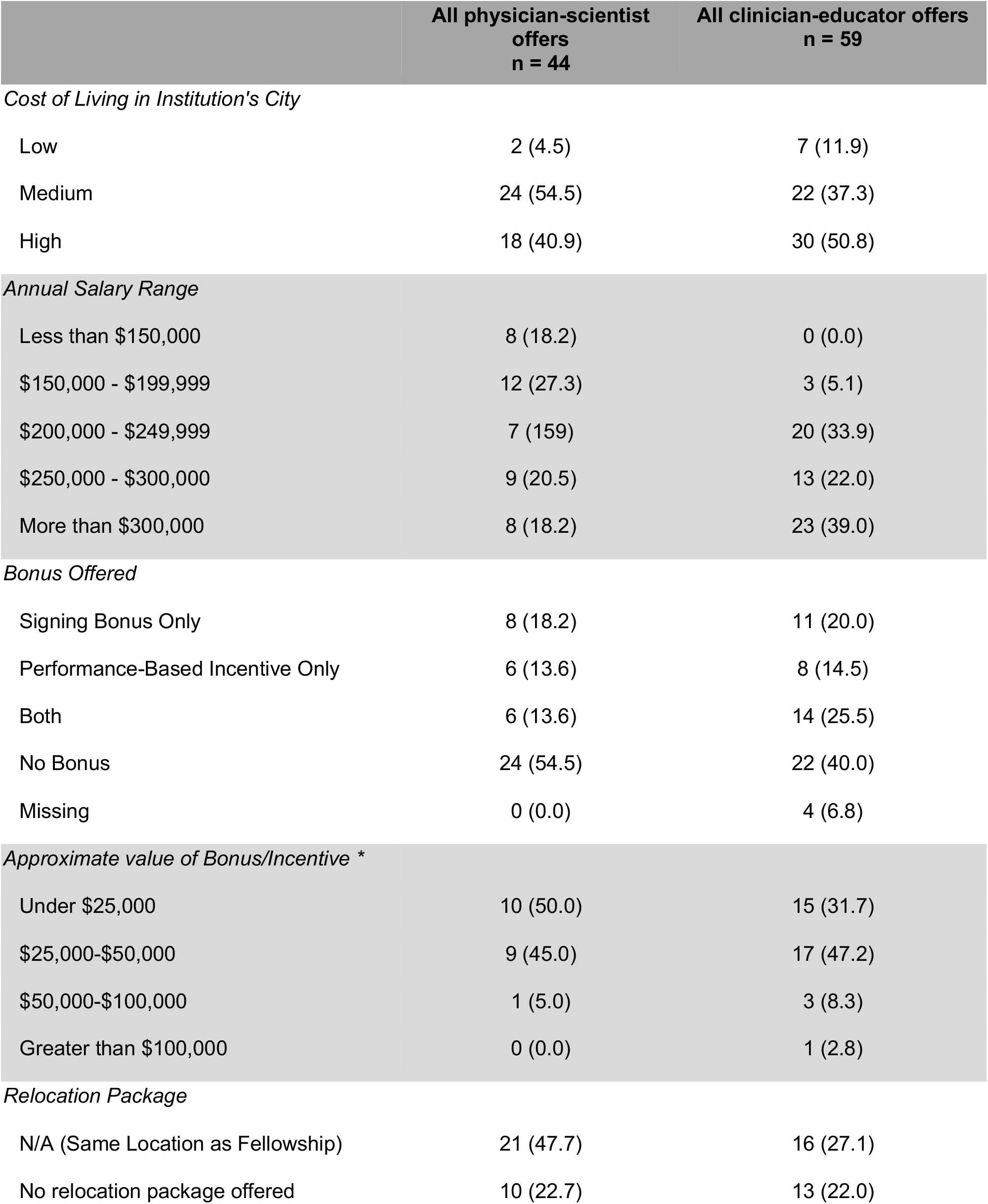
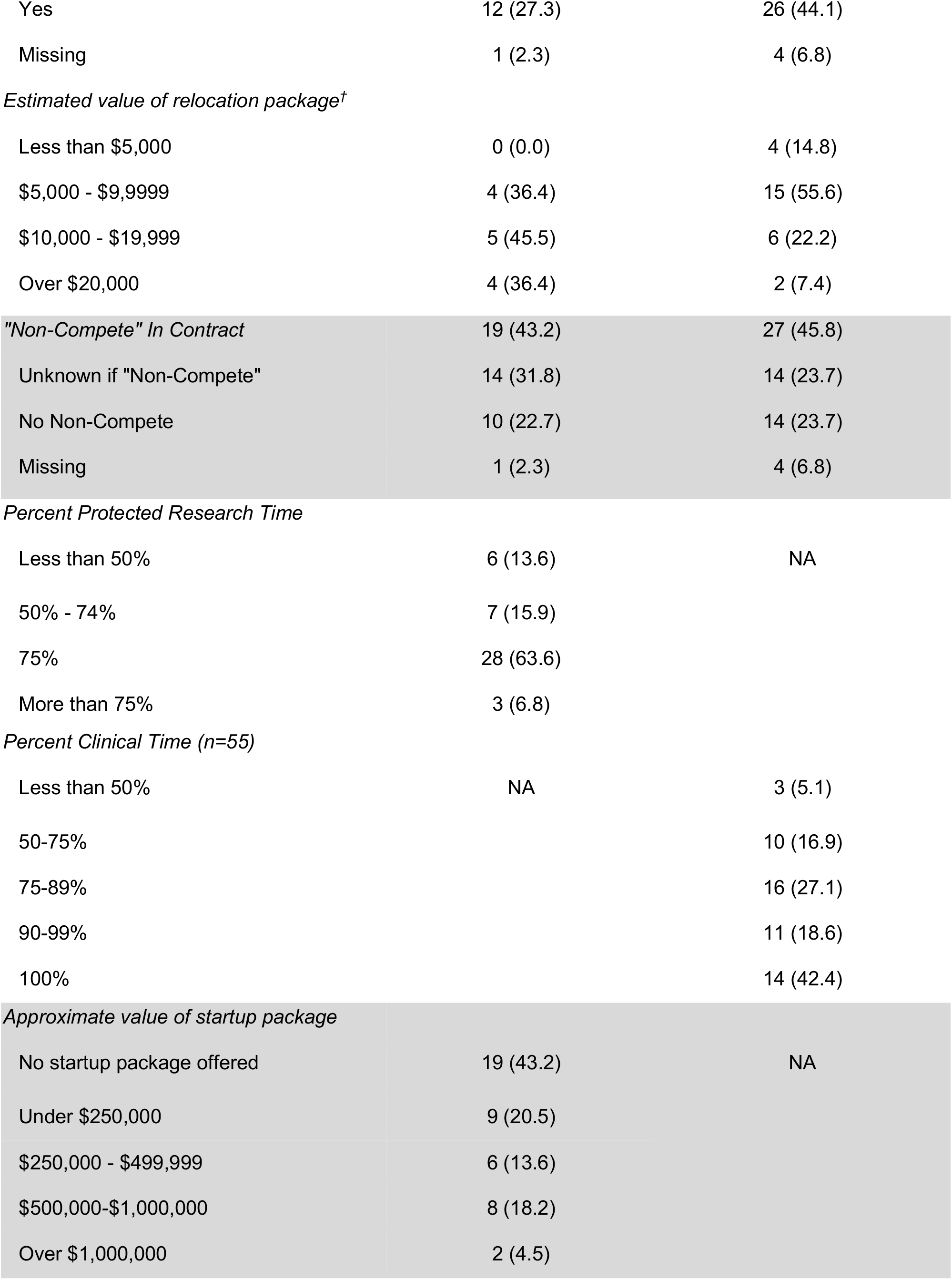
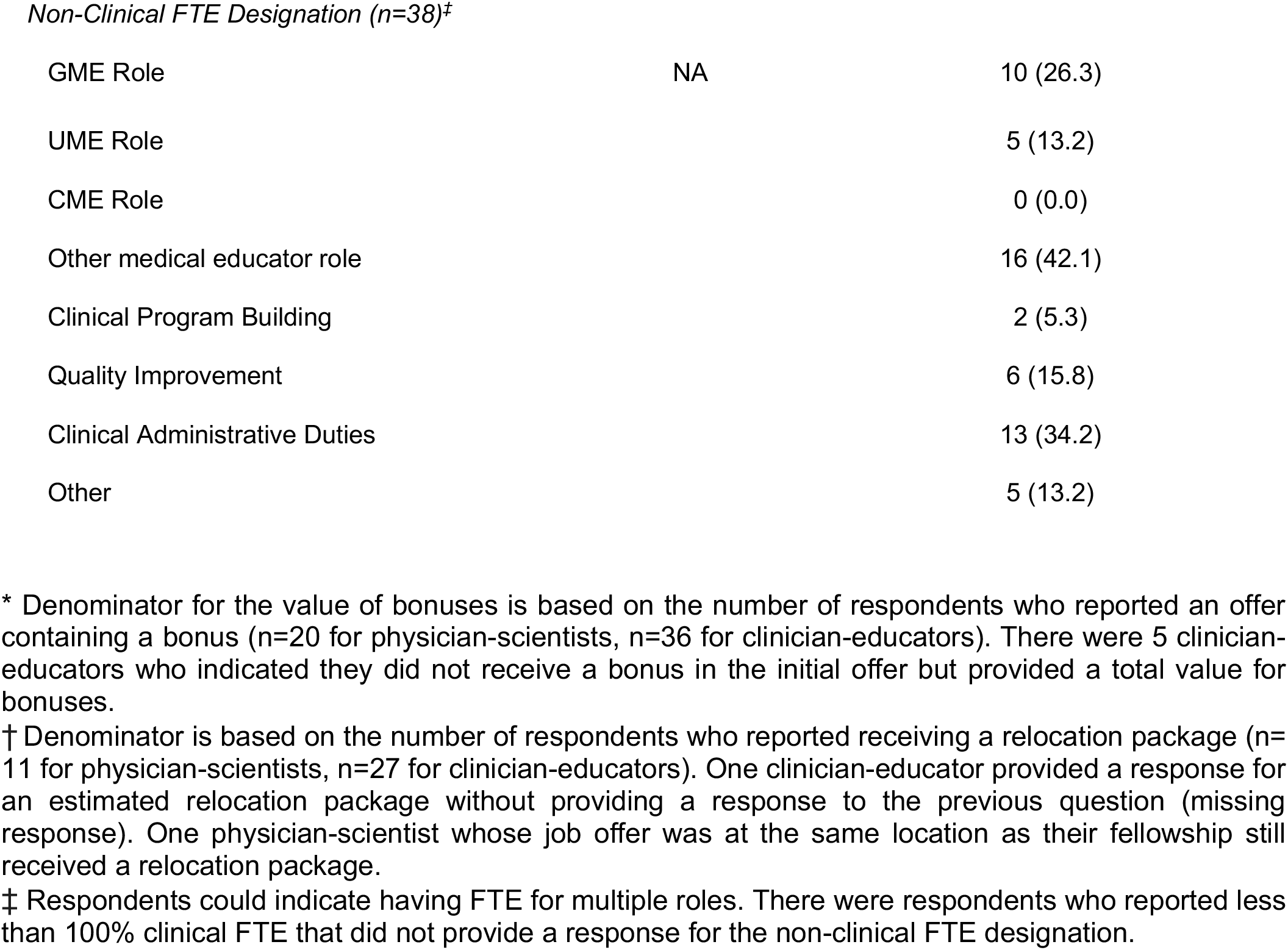
Comparison of physician-scientist and clinician-educator offers, n(%).

|  | All physician-scientist<br>offers<br>n = 44 | All clinician-educator offers<br>n = 59 |
| --- | --- | --- |
| *Cost of Living in Institution's City* |  |  |
| Low | 2 (4.5) | 7 (11.9) |
| Medium | 24 (54.5) | 22 (37.3) |
| High | 18 (40.9) | 30 (50.8) |
| *Annual Salary Range* |  |  |
| Less than \$150,000 | 8 (18.2) | 0 (0.0) |
| \$150,000 - \$199,999 | 12 (27.3) | 3 (5.1) |
| \$200,000 - \$249,999 | 7 (15.9) | 20 (33.9) |
| \$250,000 - \$300,000 | 9 (20.5) | 13 (22.0) |
| More than \$300,000 | 8 (18.2) | 23 (39.0) |
| *Bonus Offered* |  |  |
| Signing Bonus Only | 8 (18.2) | 11 (20.0) |
| Performance-Based Incentive Only | 6 (13.6) | 8 (14.5) |
| Both | 6 (13.6) | 14 (25.5) |
| No Bonus | 24 (54.5) | 22 (40.0) |
| Missing | 0 (0.0) | 4 (6.8) |
| *Approximate value of Bonus/Incentive \** |  |  |
| Under \$25,000 | 10 (50.0) | 15 (31.7) |
| \$25,000-\$50,000 | 9 (45.0) | 17 (47.2) |
| \$50,000-\$100,000 | 1 (5.0) | 3 (8.3) |
| Greater than \$100,000 | 0 (0.0) | 1 (2.8) |
| *Relocation Package* |  |  |
| N/A (Same Location as Fellowship) | 21 (47.7) | 16 (27.1) |
| No relocation package offered | 10 (22.7) | 13 (22.0) |
| Yes | 12 (27.3) | 26 (44.1) |
| Missing | 1 (2.3) | 4 (6.8) |
| *Estimated value of relocation package<sup>†</sup>* |  |  |
| Less than \$5,000 | 0 (0.0) | 4 (14.8) |
| \$5,000 - \$9,999 | 4 (36.4) | 15 (55.6) |
| \$10,000 - \$19,999 | 5 (45.5) | 6 (22.2) |
| Over \$20,000 | 4 (36.4) | 2 (7.4) |
| *"Non-Compete" In Contract* | 19 (43.2) | 27 (45.8) |
| Unknown if "Non-Compete" | 14 (31.8) | 14 (23.7) |
| No Non-Compete | 10 (22.7) | 14 (23.7) |
| Missing | 1 (2.3) | 4 (6.8) |
| *Percent Protected Research Time* |  |  |
| Less than 50% | 6 (13.6) | NA |
| 50% - 74% | 7 (15.9) |  |
| 75% | 28 (63.6) |  |
| More than 75% | 3 (6.8) |  |
| *Percent Clinical Time (n=55)* |  |  |
| Less than 50% | NA | 3 (5.1) |
| 50-75% |  | 10 (16.9) |
| 75-89% |  | 16 (27.1) |
| 90-99% |  | 11 (18.6) |
| 100% |  | 14 (42.4) |
| *Approximate value of startup package* |  |  |
| No startup package offered | 19 (43.2) | NA |
| Under \$250,000 | 9 (20.5) | |
| \$250,000 - \$499,999 | 6 (13.6) | |
| \$500,000-\$1,000,000 | 8 (18.2) | |
| Over \$1,000,000 | 2 (4.5) | |
| *Non-Clinical FTE Designation (n=38)<sup>‡</sup>* |  |  |
| GME Role | NA | 10 (26.3) |
| UME Role |  | 5 (13.2) |
| CME Role |  | 0 (0.0) |
| Other medical educator role |  | 16 (42.1) |
| Clinical Program Building |  | 2 (5.3) |
| Quality Improvement |  | 6 (15.8) |
| Clinical Administrative Duties |  | 13 (34.2) |
| Other |  | 5 (13.2) |

\* Denominator for the value of bonuses is based on the number of respondents who reported an offer containing a bonus (n=20 for physician-scientists, n=36 for clinician-educators). There were 5 clinician-educators who indicated they did not receive a bonus in the initial offer but provided a total value for bonuses.
† Denominator is based on the number of respondents who reported receiving a relocation package (n=11 for physician-scientists, n=27 for clinician-educators). One clinician-educator provided a response for an estimated relocation package without providing a response to the previous question (missing response). One physician-scientist whose job offer was at the same location as their fellowship still received a relocation package.
‡ Respondents could indicate having FTE for multiple roles. There were respondents who reported less than 100% clinical FTE that did not provide a response for the non-clinical FTE designation.

**Figure 2:**
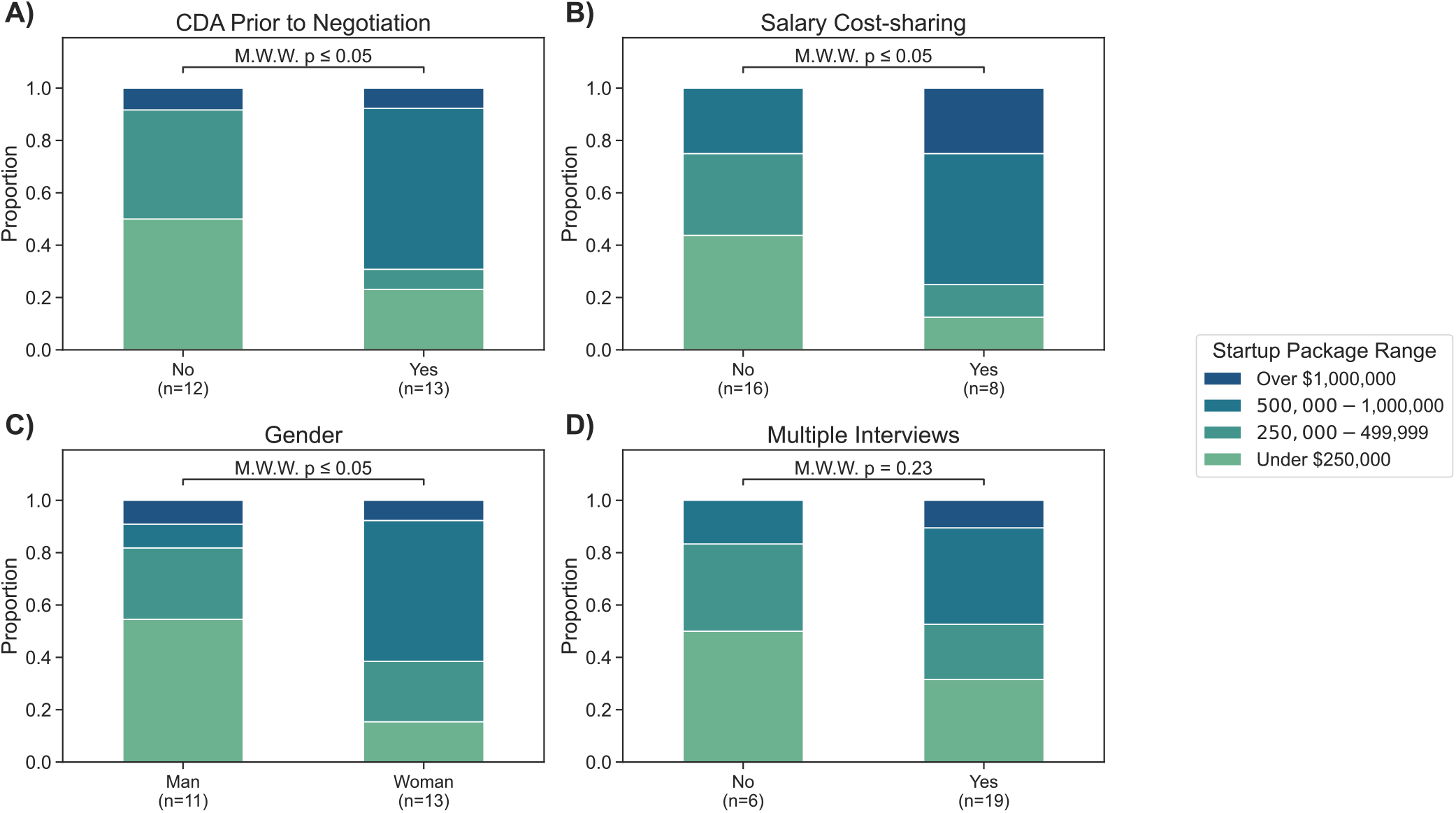
Proportional distribution of startup package ranges for physician-scientists shown as stacked bar charts. Sample sizes are indicated below each bar. Statistical comparisons performed using Mann-Whitney Wilcoxon (M.W.W.) test with p-values shown above bars. (A) CDA prior to negotiation comparison. (B) Salary cost-sharing. (C) Gender (Man vs. Woman). (D) Recipients of multiple offers versus single offer.

For all clinician-educator job offers (n=59) (**Table 2**), 42.4% had non-clinical FTE in the initial offer. Ten percent (6/56) had to negotiate for this time, whereas 35.6% indicated the non-clinical FTE was part of the initial offer and 1.7% (1/56) had external funding for the FTE. Non-clinical FTE was designated for a variety of roles. Ten percent (6/55) indicated having FTE for research in an offer. There were no significant differences for clinical FTE when comparing by gender, those who had multiple interviews, those who had an additional degree or internal/external offers (**Figure 3**). For those who have been in their role for more than a year, 77% (17/22) had a change in their FTE distribution.

**Figure 3:**
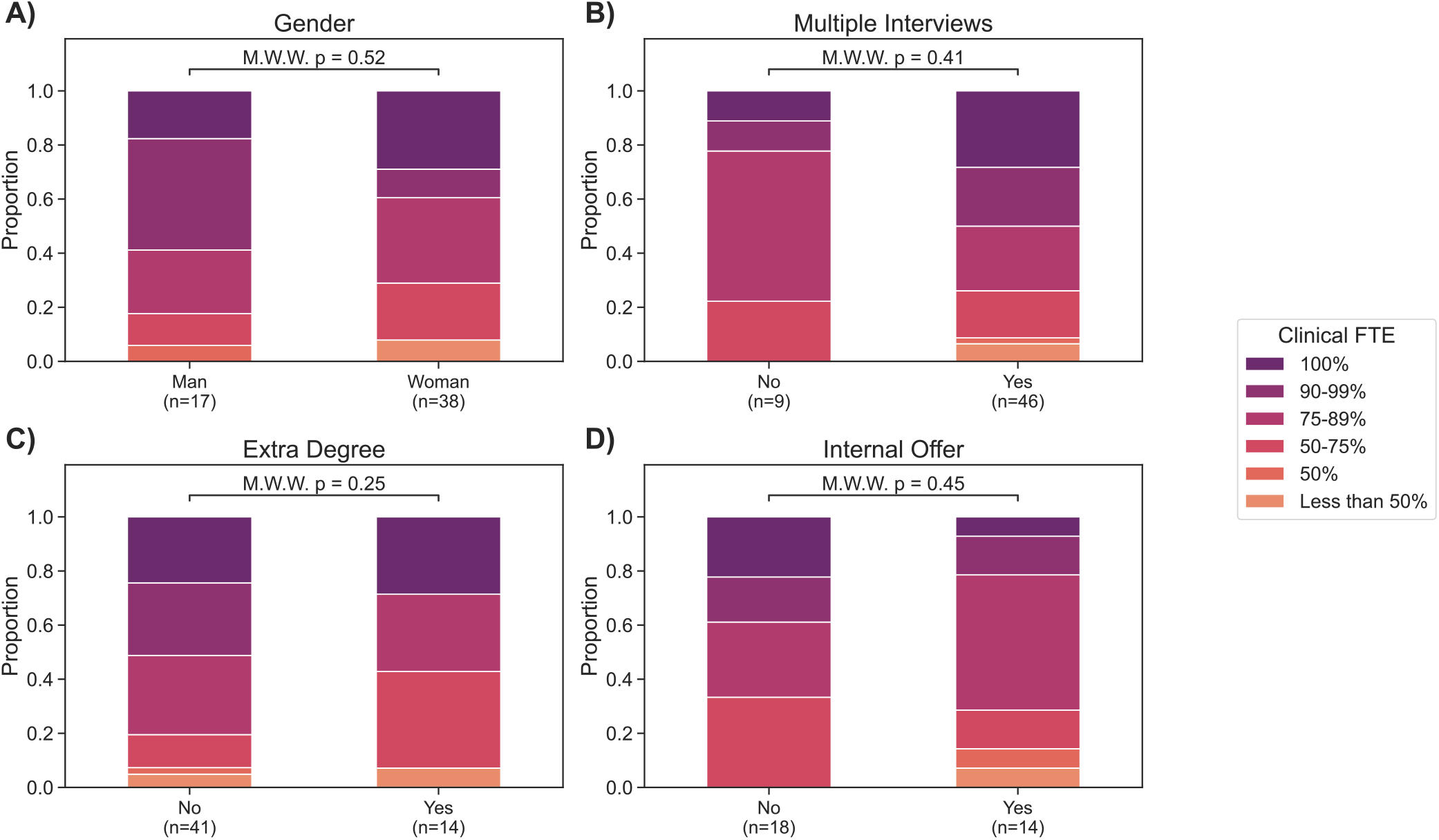
Proportional distribution of percent clinical FTE for clinician-educators as stacked bar charts. Sample sizes are indicated below each bar. Statistical comparisons performed using Mann-Whitney Wilcoxon (M.W.W.) test with p-values shown above bars. Stratified by: (A) Gender (Man vs. Woman). (B) Recipients of multiple offers versus single offer. (C) Physicians with additional advanced degrees (MS, PhD, etc.) versus MD only. and (D) Internal versus external job offers.

Ten physician-scientists wrote free-text comments about the job offer they accepted, with two including additional comments about offers they chose not to accept. Eighteen clinician-educators wrote free-text comments about the job offer they accepted, with four including additional comments about offers they chose not to accept. Free text comments about the negotiation process fell within the Social Cognitive Career Theory framework,(28) focusing on self-efficacy, outcomes, and personal goals (**Supplemental Table 2**).

Eight respondents commented they did not have negotiating power, highlighting challenges with self-efficacy. Many noted limitations particularly if staying at their home institution:

- “I felt totally trapped at my home institution since there are no other local places with [health science research] infrastructure and mentorship, and therefore also had no negotiation power.”
- “I was told that the trade-off of staying where you did fellowship was that you didn’t have to move but you had no leverage for a start-up. I got a computer and a desk as a start-up and that was it.”
- “I felt negotiation wasn’t an option.”

Additionally, several clinician-educators noted turning down offers in which moonlighting was a job expectation.

Other comments revealed both physician-scientists and clinician-educators weighed competing personal goals (family needs, geography, salary) and job outcomes (prospects for career growth). Physician-scientists most commonly commented they accepted a position due to prospects of research success (n=5), followed by family needs (n=2). For clinician-educators, the most common reasons for accepting a position were family needs (n=8) and prospects for career growth (n=8), with protected non-clinical time often being explicitly named as attractive.

## Discussion

To our knowledge, this survey is the first to report on the wide variation in reported financial support mechanisms for the first academic job among PCCM physicians. Most notably, there were statistically significant differences in payment structures for physician-scientists and clinician-educators. These differences are reflected in the non-financial aspects of a job. Physician-scientists generally give up some direct compensation to receive startup funding and highly protected time; clinician-educators receive more direct compensation, but in exchange have less flexibility for protected non-clinical time. In qualitative descriptions of the negotiation process, we identified early career physicians put value in other non-financial aspects like family and mentorship. Job contracts are highly individualized, and by describing salary ranges, bonuses, startup packages, and FTE, we hope PCCM trainees can better understand the current landscape of academic medicine as they enter negotiation discussions.

Physician-scientists responding to our survey highlighted that while having a funded CDA in-hand was not associated with salary, it was associated with higher startup packages. Although mentorship and additional education(1, 4, 29) have been shown to bolster career development as physician-scientists, financial support is an essential aspect of advancing one’s career. Prior studies identified gender differences in salary and funding, but we did not observe women receiving lower offers than men.(9, 10, 30) However, other forms of gender inequity occur throughout physician-scientist training,(9, 10, 31) which were not explored in our study. Additional methods to ensure equity beyond salary and startup funding should be considered.

For clinician-educators, we did not find statistically significant differences in clinical FTE across several comparisons. The definition of “academic time” (i.e. non-clinical FTE) is highly variable by institution.(32) Furthermore, the amount of FTE support for certain educator roles is vastly different.(33) As institutions continue re-defining clinical educator promotion pathways, there will be a simultaneous call to better describe what constitutes FTE.(34) Interestingly, the majority had adjustments to their FTE since starting. Prior research highlights gender discrimination and lack of adequate mentorship as a reason physicians pursue careers outside of academia.(8, 35) However, we found no gender differences in clinical FTE offered.

As institutions critically assess the future of academic medicine, they must continue building systems to address other aspects of systemic bias. The low number of survey respondents identifying as underrepresented in medicine limits any specific analysis. Given decreasing matriculation of applicants identifying as underrepresented into PCCM fellowship, this highlights the importance of providing appropriate guidance to all to maintain a diverse workforce.(36, 37) Trainees must also understand funding structures to better navigate negotiation conversations.(19) As highlighted in free text responses, there are many important aspects besides salary to job negotiation and offer acceptance, including family and location. Future research should seek to better understand the impact of other non-salary financial benefits, including loan repayment structures, retirement contributions, childcare support, scholarships, etc.

Our study has several limitations. Importantly, the definition of academic PCCM faculty roles is relatively broad and purposely not defined as part of our survey.(32) Physicians practicing in community hospitals where there are trainees (e.g. medical students or residents) as well as with advanced training in sleep or interventional pulmonology, are likely included in the responses. Similarly, we did not delineate the type of clinical time, which may impact salary (e.g. critical care, procedure services, clinic, etc.) Respondents were likely impacted by recall bias. The survey did not capture the timing of formal job offer letters, which may have impacted negotiation discussions. Snowball sampling methodology and the small sample size limits our analysis. Based on only 20% of graduating PCCM fellows entering academic medicine, we anticipate a total of 720 eligible PCCM faculty starting academic jobs between 2020-2025.(47) However, our own networks are limited and we are unable to characterize non-respondents. Since our inclusion criteria included faculty who received job offers during the COVID pandemic, it is unclear if certain aspects of job offers were impacted.

Our exploratory survey provides informative details about the current job market in PCCM. Our results may help fellowship graduates better understand the key factors of their first job negotiation, especially since several respondents commented about their lack of negotiating power. Improved data collection and reporting on job contract features from national professional organizations like the AAMC could support the future physician workforce. Transparency about financial and non-financial components of job contracts is more about highlighting options available. Importantly, there are many intangible components to accepting a job offer that were not specifically addressed in our survey. Additional research is needed to better understand if certain aspects of the initial contract are helpful in retaining physicians in academic medicine.

## Conclusions

In a survey of 2020-2025 PCCM graduates who accepted jobs in academic medicine, there is variability in the salary range between physician-scientists and clinician-educators. Physician-scientists having a funded CDA such as a K23 significantly impacted their start-up package. Clinician educators had wide ranges in non-clinical FTE and salary offers. We hope details from our survey will help future academics navigate their first job search.

## Data Availability

Code availability: Code for our analysis is available at https://github.com/cag-lab/pccm_job_negotiation.
Data availability: The structured data have been deposited to the Harvard Dataverse at https://doi.org/10.7910/DVN/NI3K31 and are available to researchers who agree to outlined terms and complete a data request form.

## Abbreviations

PCCM: Pulmonary and Critical Care Medicine
FTE: Full Time Equivalent
CDA: Career Development Award
AAMC: Association of American Medical Colleges
AMA: American Medical Association

## Code availability

Code for our analysis is available at https://github.com/cag-lab/pccm_job_negotiation.

## Data availability

The structured data have been deposited to the Harvard Dataverse at https://doi.org/10.7910/DVN/NI3K31 and are available to researchers who agree to outlined terms and complete a data request form.

## Acknowledgements

We are grateful to our colleagues willing to share their experiences with the negotiation process openly. We would like to thank Joe A Hippensteel, Isadore M Budnick, Priscilla De La Cruz, and Scott A Laurenzo for feedback during survey development. We would like to thank Matt Christensen, Abigail Chua, and PulmPEEPs (David Furfaro and Kristina Montemayor) for further sharing the survey. We would like to thank David Liebovitz and Nikolay S Markov for coding/analysis/visualization feedback.

## DATA SUPPLEMENT

## Supplemental Material 1. Survey (see separate file)

## Supplemental Data 1: Incomplete data

Data incomplete across job offers (n=103):

Gender: 1 (0.9%) selected ‘Prefer not to answer’ and not included in gender-based analyses Salary: 9 (8.7%)

Bonuses: 4 (0.97%)

Bonus value: 47 (45.6%)

Clinician educators job offers (n=59)

Non-clinical FTE: 3 (5.1%)

FTE for research: 4 (6.8%)

Salary adjusted based on clinical effort: 4 (6.8%) Salary equity transparency: 4 (6.8%)

Relocation package: 4 (6.8%)

Physician-scientist job offers (n=44)

Startup package value: 19 (43.2%) – some of whom answered ‘no’ to receiving a startup package

Startup cost sharing: 16 (36.4%) – some of whom answered ‘no’ to receiving a startup package

Relocation package: 1 (2.3%)

Non-complete clause: 1 (2.3%)

**Supplemental Table 1:** Free text descriptions of non-compete clauses in contracts

|  |  |
| --- | --- |
| Description of Non-Compete Clauses for both physician-scientist and clinician-educator jobs | Most did not provide additional details. Those who did described various aspects:<br>- Time ranged from 1 - 2 years<br>- Location ranged from 3-50 miles from primary site, 25 miles from any credentialed site, entire county or even entire state<br>- Buyout ranged from 0 - 50k<br>- Some commented on a non-compete for moonlighting in other hospitals<br>- "I probably signed a non-compete but do not remember it" |

**Supplemental Table 2:** Social Cognitive Career Theory categorization of free text with representative quotes

|  |  |
| --- | --- |
| Social Cognitive Career Theory Domain | Representative quote |
| Self-efficacy | - "Used competing offer to increase salary from 125 to 135k in negotiation" - physician-scientist<br>- "It was a clinical instructor pathway that had only one contract pipe and payment structure - two years to get grant funding. No negotiation was allowed... in retrospect would not recommend such a pathway for anyone in the future." - physician-scientist<br>- "Despite having several best alternatives to a negotiated agreement and an extremely unique career path, I was able to negotiate very little compared to the starting offer coming from the institution that I ultimately took the job at." - physician-scientist<br>- "I was told that the trade-off of staying where you did fellowship was that you didn't have to move but you had no leverage for a start-up. I got a computer and a desk as a start-up and that was it." - physician-scientist<br>- "My job provided the best of both worlds- I work with trainees without having the burden that comes with academic medicine. Additionally, my job is not RVU based, it's salaried, so I do not need to stress about meeting certain goals and I can provide high-level medical care without worrying about income." - clinician-educator<br>- "Low salary [but] institutional familiarity, and really needed to be in the location." - clinician-educator<br>- "Salary support would have to come from 'nocturnal moonlighting' to get critical care time. Academic productivity expected to come out of faculty's own time and support (grants, non-clinical FTE, etc.) built up after initial salary protection." - clinician-educator<br>- "It was the lowest pay option with minimal to no chance of upward mobility or pay increase since I was not going to become a researcher." - clinician-educator |
| Outcome expectations | - "Better start-up package, more opportunities to expand research, higher salary, assistant professor instead of instructor" - physician-scientist<br>- "More protected research time" - physician-scientist<br>- "I felt totally trapped at my home institution since there are no other local places with [health science research] infrastructure and mentorship, and therefore also had no negotiation power" - physician-scientist<br>- "I have a mentor in what I'm interested in doing in my career." - clinician-educator<br>- Clinician-educators frequently commented about:<br>- Future GME opportunities<br>- Protected education time |
| Personal goals | - "Family" or "geography" was frequently mentioned for both physician-scientists and clinician-educators. For example:<br>- "Prioritized being close to family; not our best job offer by pay/time etc, but in city with family." - clinician-educator<br>- "Not much room for negotiation. Extra stipend given for clinical director title. We did have non clinical time built in to use for research, clinical work etc even with contract as 'full time clinical'." |
| | - clinician-educator<br>- "Additional \$2k monthly fellowship stipend from time of signing to start date" - clinician-educator |

## References

1. Borges NJ, Navarro AM, Grover A, Hoban JD. How, when, and why do physicians choose careers in academic medicine? A literature review. Acad Med 2010;85:680–686.

2. Herchline D, Cohen ME, Ambrose M, Hwang J, Kaminstein D, Kilberg M, et al. Into the Unknown: Characterizing Fellow Uncertainty During the Transition to Unsupervised Practice. J Grad Med Educ 2023;15:201–208.

3. Nonnemaker L. Women physicians in academic medicine: new insights from cohort studies. N Engl J Med 2000;342:399–405.

4. Nadig NR, Vanderbilt AA, Ford DW, Schnapp LM, Pastis NJ. Variability in Structure of University Pulmonary/Critical Care Fellowships and Retention of Fellows in Academic Medicine. Annals of the American Thoracic Society 2015;12:553.

5. Kubiak NT, Guidot DM, Trimm RF, Kamen DL, Roman J. Recruitment and retention in academic medicine--what junior faculty and trainees want department chairs to know. Am J Med Sci 2012;344:24–27.

6. Weinert CR, Billings J, Ryan R, Ingbar DH. Academic and career development of pulmonary and critical care physician-scientists. Am J Respir Crit Care Med 2006;173:23–31.

7. Bates C, Gordon L, Travis E, Chatterjee A, Chaudron L, Fivush B, et al. Striving for Gender Equity in Academic Medicine Careers: A Call to Action. Acad Med 2016;91:1050–1052.

8. Catenaccio E, Rochlin JM, Simon HK. Addressing Gender-Based Disparities in Earning Potential in Academic Medicine. JAMA Netw Open 2022;5:e220067.

9. Jagsi R, Griffith KA, Stewart A, Sambuco D, DeCastro R, Ubel PA. Gender differences in salary in a recent cohort of early-career physician-researchers. Academic medicine : journal of the Association of American Medical Colleges 2013;88:.

10. Jagsi R, Griffith KA, Stewart A, Sambuco D, DeCastro R, Ubel PA. Gender differences in the salaries of physician researchers. JAMA 2012;307:.

11. Holliday E, Griffith KA, De Castro R, Stewart A, Ubel P, Jagsi R. Gender differences in resources and negotiation among highly motivated physician-scientists. J Gen Intern Med 2015;30:401–407.

12. AAMC Faculty Salary Report. AAMC at <https://www.aamc.org/data-reports/workforce/report/aamc-faculty-salary-report>.

13. Doximity. Doximity 2024 Physician Compensation Report. At <https://www.doximity.com/reports/physician-compensation-report/2024>.

14. Critical Care Specialists Salary in Chicago, IL. at <https://www.medscape.com/physician-salary-explorer?specialty=Critical%20Care&city=Chicago&state=IL&dma=Chicago&region=Great%20Lakes&yoe=1%2C7&emp=1>.

15. Baimas-George M, Fleischer B, Korndorffer JR, Slakey D, DuCoin C. The Economics of Private Practice versus Academia in Surgery. Journal of surgical education 2018;75:.

16. Mason BS, Landry A, Sánchez JP, Williams VN. How to Find an Academic Position After Residency: Who, What, When, Where, Why, and How. MedEdPORTAL 2018;14:10727.

17. Oberle AJ, Kumar S, Nelson M, Kraft M, Lenz PH. Searching for the First Job: A Practical Guide for Fellows-in-Training. Chest 2019;155:25–32.

18. Cerimele JM, Bauer AM, Ratzliff A. Guiding Academic Clinician Educators at Research-Intensive Institutions: a Framework for Chairs, Chiefs, and Mentors. J Gen Intern Med 2022;

19. Chang A, Schwartz BS, Harleman E, Johnson M, Walter LC, Fernandez A. Guiding Academic Clinician Educators at Research-Intensive Institutions: a Framework for Chairs, Chiefs, and Mentors. J Gen Intern Med 2021;36:3113–3121.

20. Berman RA, Gottlieb AS. Job Negotiations in Academic Medicine: Building a Competency-Based Roadmap for Residents and Fellows. Journal of General Internal Medicine 2018;34:146.

21. Sambuco D, Dabrowska A, Decastro R, Stewart A, Ubel PA, Jagsi R. Negotiation in academic medicine: narratives of faculty researchers and their mentors. Acad Med 2013;88:505–511.

22. Gilbert J, Kothari P, Sanchez N, Spencer DJ, Soto-Greene M, Sánchez JP. Is Academic Medicine a Financially Viable Career? Exploring Financial Considerations and Resources. MedEdPORTAL 2020;16:10958.

23. Hochberg CH, Eakin MN. Keys to successful survey research in health professions education. ATS Sch 2024;5:206–217.

24. Harris PA, Taylor R, Thielke R, Payne J, Gonzalez N, Conde JG. Research electronic data capture (REDCap)--a metadata-driven methodology and workflow process for providing translational research informatics support. J Biomed Inform 2009;42:377–381.

25. Valerio MA, Rodriguez N, Winkler P, Lopez J, Dennison M, Liang Y, et al. Comparing two sampling methods to engage hard-to-reach communities in research priority setting. BMC Med Res Methodol 2016;16:146.

26. Methangkool E, Brodt J, Kolarczyk L, Ivascu NS, Hicks MH, Herrera E, et al. Perceptions of Gender Disparities Among Women in Cardiothoracic Anesthesiology. J Cardiothorac Vasc Anesth 2022;36:1859–1866.

27. Social Cognitive Career Theory. doi:10.1016/B978-0-12-818630-5.13022-2.

28. Patton W, McMahon M. Career Development and Systems Theory: Connecting Theory and Practice (4th Edition). BRILL; 2021.

29. Panettieri RA Jr, Kolls JK, Lazarus S, Corder S, Harshman A, Langmack E, et al. Impact of a Respiratory Disease Young Investigators’ Forum on the Career Development of Physician-Scientists. ATS Sch 2020;1:243–259.

30. Sege R, Nykiel-Bub L, Selk S. Sex Differences in Institutional Support for Junior Biomedical Researchers. JAMA 2015;314:1175–1177.

31. Ghosh-Choudhary S, Carleton N, Flynn JL, Kliment CR. Strategies for Achieving Gender Equity and Work-Life Integration in Physician-Scientist Training. Acad Med 2022;97:492–496.

32. Misra M, Huang GC, Becker AE, Bates CK. Leaders’ Perspectives on Resources for Academic Success: Defining Clinical Effort, Academic Time, and Faculty Support. Perm J 2024;28:33–41.

33. Glod SA, Alexandraki I, Jasti H, Lai CJ, Ratcliffe TA, Walsh K, et al. Clerkship Roles and Responsibilities in a Rapidly Changing Landscape: a National Survey of Internal Medicine Clerkship Directors. J Gen Intern Med 2020;35:1375–1381.

34. Block SM, Sonnino RE, Bellini L. Defining “faculty” in academic medicine: responding to the challenges of a changing environment. Acad Med 2015;90:279–282.

35. Edmunds LD, Ovseiko PV, Shepperd S, Greenhalgh T, Frith P, Roberts NW, et al. Why do women choose or reject careers in academic medicine? A narrative review of empirical evidence. Lancet 2016;388:2948–2958.

36. Sánchez JP. Succeeding in Academic Medicine: A Roadmap for Diverse Medical Students and Residents. Springer Nature; 2020.

37. Santhosh L, Babik JM. Diversity in the Pulmonary and Critical Care Medicine Pipeline. Trends in Gender, Race, and Ethnicity among Applicants and Fellows. ATS Sch 2020;1:152–160.

38. Strumpf Z, Miller C, Abbas KZ, Bensken WP, Matta M. Trends in Pulmonary Critical Care Fellowship Applications and Match Rates before and after the Onset of the COVID-19 Pandemic. ATS Scholar 2024; doi:10.34197/ats-scholar.2023-0057OC.

